# Mammary epithelium permeability during established lactation: Associations with cytokine levels in human milk

**DOI:** 10.1101/2022.11.18.22282518

**Authors:** Katie T. Kivlighan, Sallie S. Schneider, Eva P. Browne, Brian T. Pentecost, Douglas L. Anderton, Kathleen F. Arcaro

## Abstract

**Objective:** The cytokine profile of human milk may be a key indicator of mammary gland health and has been linked to infant nutrition, growth, and immune system development. The current study examines the extent to which mammary epithelium permeability (MEP) is associated with cytokine profiles during established lactation among a diverse group of US mothers.

**Methods:** Participants were drawn from a previous study of human milk cytokines. The present analysis includes 162 participants (98 Black women, 64 White women) with infants ranging from 1-18 months of age. Levels of cytokines were determined previously. Here we measure milk sodium (Na) and potassium (K) levels with ion-selective probes. *Elevated MEP* was defined and evaluated by both Na levels > 10 mmol/L and Na/K ratios greater than 0.6. Bivariate associations, principal components analysis, and multivariable logistic regression models were used to examine associations between maternal-infant characteristics, twelve cytokines (IL-6, IL8, TNFα, IL-1β, FASL, VEGFD, FLT1, bFGF, PLGF, EGF, leptin, adiponectin), and elevated MEP.

**Results:** Elevated MEP was observed in 12% and 15% of milk samples as defined by Na and Na/K cutoffs, respectively. The odds of experiencing elevated MEP (defined by Na > 10 mmol/L) were higher among Black participants and declined with older infant age. All cytokines, except leptin, were positively correlated with either Na or the Na/K ratio. A pro-inflammatory factor (IL-6, IL-8, TNFα, IL-1β, EGF) was more strongly correlated with the Na/K ratio, while a tissue remodeling factor (FASL, VEGFD, FLT1, bFGF, PLGF, adiponectin) was more strongly correlated with Na. The pro-inflammatory and tissue remodeling factors each contributed uniquely to raising the odds of elevated MEP as defined by either Na or the Na/K ratio.

**Conclusion:** This exploratory analysis of MEP and cytokine levels during established lactation indicates that elevated MEP among US women may be more common than previously appreciated and that Black women may have increased odds of experiencing elevated MEP based on current definitions. Research aimed at understanding the role of MEP in mammary gland health or infant growth and development should be prioritized.

## 1 Introduction

Human milk is a complex biological fluid containing a multitude of cellular and molecular components integral to the health or disease state of the mother and infant. In particular, inflammatory markers in human milk appear to have a role in regulating infant nutrition (1,2), growth (3,4), and immune system development (5,6), and may be useful indicators of current (7,8) and future mammary gland health (9,10).

Human milk contains a variety of cytokines that act locally in the mammary gland and/or influence the growth and development of infant tissues (11). Cytokines represent a large class of secreted bioactive molecules that modulate cell-to-cell communication to impact cellular growth, viability, and differentiation, as well as immune and inflammatory responses (5). In human milk, cytokines are produced by leukocytes that have migrated into the mammary tissue from systemic circulation as well as by tissue-resident cells, such as mammary epithelium, fibroblasts, and adipocytes (5,12). Specific cytokines are upregulated during mammary gland differentiation during pregnancy, in response to milk stasis, and during involution (13,14). This upregulation is distinct from the rise in pro-inflammatory cytokines observed during an infection, such as mastitis. In the setting of infection, pro-inflammatory cytokines modulate the immune response of the mammary gland (7,8).

Adipokines, such as leptin and adiponectin, are a subset of cytokines produced by adipose tissue that serve as endocrine signaling molecules with roles in regulating metabolism and body composition (11,12). Both also play an important role in modulating mammary gland development and tissue remodeling during the lactation cycle. Growth factors are a class of cytokines linked to tissue growth and remodeling. They may retain bioactivity after ingestion and are important for the development of the infant intestinal barrier (11). Within the mammary gland, growth factors are important for angiogenesis, maintaining lactation, and regulating involution (15,16).

Measurement of mammary epithelium permeability (MEP) may provide important information for the interpretation of cytokine concentrations in human milk. Prior to secretory activation (i.e., onset of mature milk), paracellular pathways between mammary epithelial cells are open allowing communication between the maternal bloodstream and the mammary gland (17). Following birth, rapid tight junction formation within the mammary epithelium facilitates paracellular pathway closure, a shift important for the establishment and maintenance of milk synthesis and secretion (18,19). The paracellular pathway is particularly important for the movement of ions across the mammary epithelium. For example, sodium (Na) levels are high in colostrum, but decrease rapidly during the first 5 days postpartum in response to paracellular pathway closure. It has been proposed that once Na levels decrease below 10 mmol/L, milk maturity has been achieved (17,20). In contrast, potassium (K) accumulates in milk as paracellular pathways close (17,21). Both Na alone and as a ratio with potassium (Na/K) have been used to assess MEP. Na/K ratios of less than 0.6 have been used to indicate tight junction closure and milk maturity (17).

As long as the paracellular pathways remain closed during established lactation, milk secretion is maintained and levels of Na and the Na/K ratio remain low (18). However, both milk accumulation and inflammation have been linked to the re-opening of tight junctions and rising Na or Na/K ratios (18,22). Our lab recently demonstrated that shifts in the Na/K ratio were closely linked to anti-SARS-CoV-2 antibody levels in human milk within a single individual over time, confirming an association between permeability and immune factors (25). Knowledge of MEP could provide important context for the interpretation of cytokine levels in human milk.

We previously examined levels of human milk cytokines in a diverse cohort of women during established lactation to determine associations with obesity, race, and risk factors for breast cancer (24). Of note, certain pro-inflammatory cytokines (IL-1β, FASL), growth factors (bFGF, EGF), and adipokines (leptin, adiponectin) were elevated among participants with a BMI >30. Levels of 1L-1β and leptin were found to be higher in Black participants.

The goals of the current study were 1) to determine the extent to which MEP as indicated by Na and Na/K ratios is associated with cytokine profiles in human milk during established lactation and 2) to determine if race or BMI are associated with elevated MEP. We hypothesized that higher levels of human milk pro-inflammatory cytokines would be associated with elevated permeability and that similar to observed patterns for the cytokines listed above, elevated MEP would be more common among participants with Black race or a BMI >30.

## 2 Methods

### 2.1 Study Population

This is a secondary analysis of selected data from a study examining racial differences in cytokines in human milk in relation to breast cancer etiology (24). For the present study, we selected participants for whom archived whole milk samples were available. All participants had signed a consent form for an IRB-approved study that included secondary sample analysis. Briefly, for the original study, all participants completed questionnaires on demographics and health history. Milk samples were collected between 2007 and 2013 from lactating women aged 18 years and older living in the continental United States (US). Participants had collected milk in the morning upon waking by expressing the full contents of each breast into separate glass or BPA-free plastic containers via hand expression or use of their own pump. Archived aliquots of whole milk used in this project had been stored at −20°C.

Of the 292 participants in the original study, frozen aliquots of whole milk were available from 167 participants (24). Of these, 5 mother-infant dyads were identified as being more than 3 SD above the mean for infant age (>798 days or 2.18 years of age). Breastfeeding practices employed by these participants were not typical of the remainder of the sample and were therefore excluded. Our final sample consisted of 162 participants (98 Black women and 64 White women) with infants ranging in age from 1 to 18 months.

### 2.2 Measurement of Sodium and Potassium Ions

Levels of Na and K were measured using ion-selective electrode probes (Medica EasyLyte Na/K Analyzer), providing new data for the current analysis. Briefly, 1 mL aliquots of whole milk were thawed, centrifuged at 3220 g for 3 minutes at 4°C and the Na and K concentrations (mmol/L) were determined in the clarified whey fraction. A total of 18 samples were run in duplicate with mean coefficients of variation (CV) of 7.8% for Na and 4.4% for K. A ratio was calculated between Na and K for each participant. Neither Na, K, nor the Na/K ratio were associated with medication use, freeze-thaw cycles, or shipping methods.

### 2.3 Measurement of Cytokines and Growth Factors

Assays for pro-inflammatory cytokines, growth factors, and adipokines were performed in the original study (24). Briefly, multiplex and single-analyte electrochemical-luminescent sandwich assays from MesoScale Discovery (MSD, Gaithersburg, MD) were used to measure the 15 analytes, of which 12 were selected for the present analysis and are shown in **Table 1**. INFγ, TIE-2, and VEGFC were excluded due to low detectability (< 35%). Thirty-eight samples, eight standards, and two of three controls were tested in duplicate on each plate according to the manufacturer’s protocols. Coefficients of variation (CVs) and intraclass correlation coefficients (ICCs) are available in Murphy et al. (24).

**Table 1.** Lower limits of detection (LLOD) for pro-inflammatory cytokines, growth factors, and adipokines in human milk from Murphy et al.(24).

|  | Analytes | Analyte Symbol | LLOD (pg/mL) |
| --- | --- | --- | --- |
| **Pro-inflammatory Cytokines** | Interleukin-6 | IL-6 | 0.14 |
|  | Interleukin-8 | IL-8 | 0.12 |
| | Tumor necrosis factor-alpha | TNFα | 0.09 |
| | Interleukin-1 beta | IL-1β | 0.07 |
| **Growth Factors** | *Fas* ligand | FASL | 0.415 |
|  | Vascular endothelial factor D | VEGFD | 3.26 |
|  | Fms-related tyrosine kinase 1 | FLT1 | 1.53 |
|  | Placental growth factor | PLGF | 0.42 |
|  | Basic fibroblast growth factor | bFGF | 0.15 |
|  | Epidermal growth factor | EGF | 0.075 |
| **Adipokines** | Leptin | Leptin | 93.96 |
|  | Adiponectin | Adiponectin | 0.08 |

Mean concentrations of human milk analytes were calculated for all 162 samples with duplicate values above the lower limit of detection (LLOD) as reported in Murphy et al. (24). The percent of mean values below the LLOD was calculated for each analyte. Values below the LLOD were imputed with the LLOD/2. Per Keizer et al. (25), single imputation with LLOD/2 has equivalent performance to maximum likelihood imputation when less than 10% of the sample is missing. For the 162 participants included in the current study, the majority of analytes (8 of 12) had less than 10% of samples below the LLOD. However, IL-6, IL-1β, TNFα, and bFGF ranged from 13% to 20% of values below the LLOD.

### 2.4 Statistical analysis

Data were cleaned and coded. All analyses were performed in SPSS 28.0. Descriptive statistics were examined. Spearman rank correlations and Mann-Whitney *U* tests were performed to explore bivariate associations. Two definitions of *elevated MEP* were examined in our dataset: Na ≥ 10 mmol/L (17,20), and Na/K ratio ≥ 0.6 (17). Multivariable logistic regression models were fitted to predict increased MEP from maternal-infant factors. Principal component analysis with varimax rotation was used to identify affinities between pro-inflammatory cytokines, growth factors, and adipokines after normalizing (natural log) and centering analytes. Based on sample size, 0.45 was designated as the threshold for factor loadings (26). Factor scores were computed based on these results. Multivariable logistic regression models were fitted to examine factor scores as predictors of elevated MEP.

## 3 Results

### 3.1 Descriptive Statistics for Na and the Na/K Ratio

Descriptive statistics for Na, K, and Na/K ratios are presented in **Table 2**. Median concentrations of Na and K were 6.2 mmol/L (range 2.8 – 23.6) and 14.6 mmol/L (range 10.1 – 28.1) respectively. The median Na/K ratio in this sample was 0.41 with a range of 0.18 to 1.08. Na and the Na/K ratio were strongly correlated, *r*(160) = 0.79, *p* < .001. Two sets of criteria for identifying elevated MEP were examined. Using the criteria defined in the literature of 10 mmol/L for Na (17,20), the cutoff for elevated MEP was at the 90^th^ percentile and a total of 19 cases of elevated MEP were identified. Using the criteria of 0.6 for the Na/K ratio (17), the cutoff for elevated MEP was at the 85^th^ percentile and 25 cases were identified. A total of 17 cases were identified by both criteria. Two cases were identified by Na ≥ 10 mmol/L alone, while 8 cases were identified solely by Na/K ≥ 0.6 (**Figure 1**).

**Figure 1.**
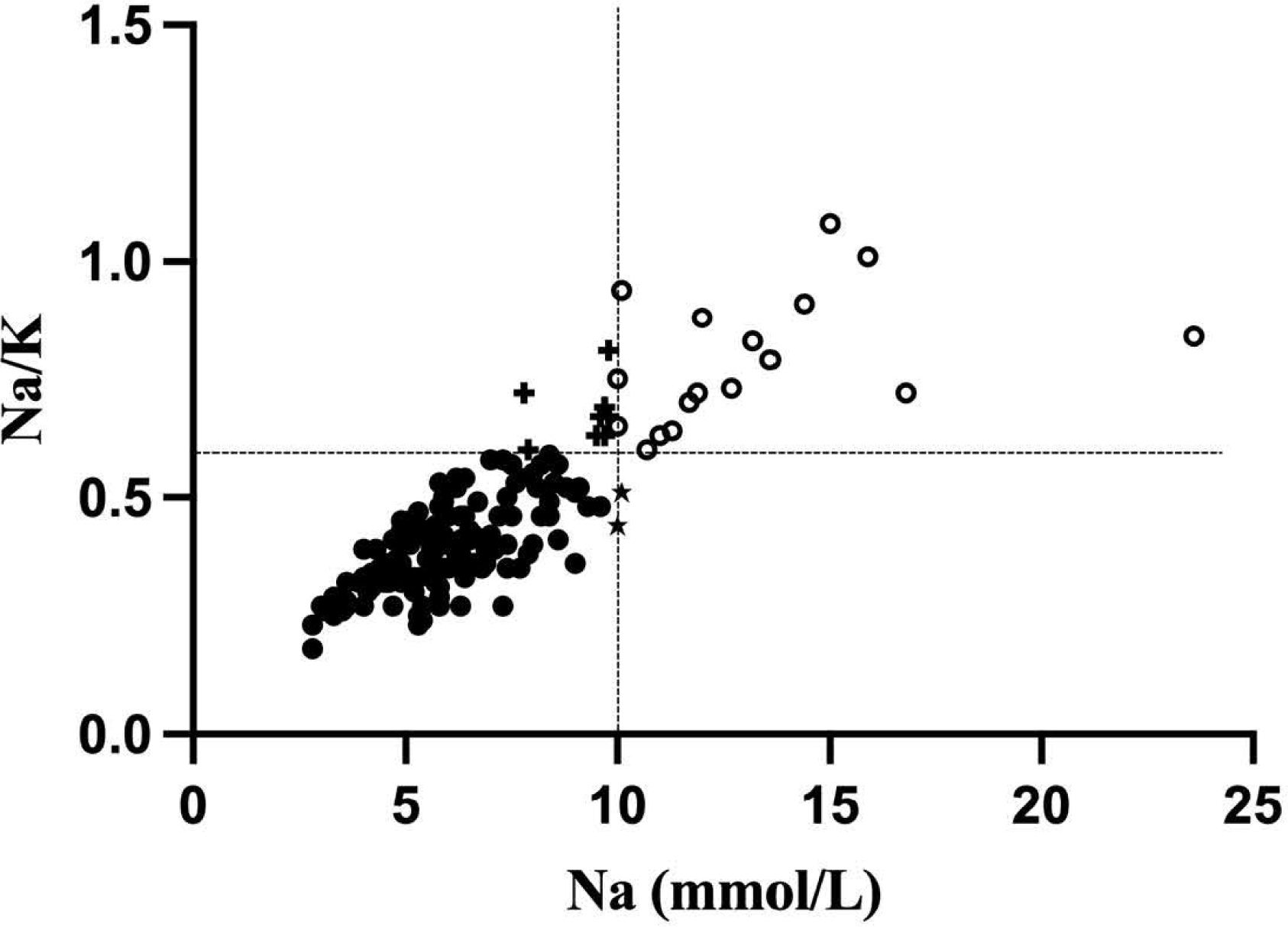
Scatterplot of sodium (Na; mmol/L) and the sodium-potassium (Na/K) ratio. Cases identified only by a Na level ≥ 10 mmol/L are indicated with a star (*n* = 2). Cases identified only by a Na/K ratio ≥ 0.6 are indicated with a plus sign (*n* = 8). Cases identified by both criteria are located in the right upper quadrant and are shown as open circles.

**Table 2.** Descriptive statistics for Na, K, and the Na/K ratio (*N* = 162) and criteria for defining elevated MEP (cutoffs bolded) (17).

|  | **Median** | **IQR** | **Elevated MEP**<br>**Criteria** |
| --- | --- | --- | --- |
| Sodium (mmol/L) | 6.2 | 5.0 – 8.0 | $\geq \mathbf{10}$ mmol/L<br>90<sup>th</sup> percentile<br>$N = 19$ |
| Potassium (mmol/L) | 14.6 | 12.7 – 17.4 | N/A |
| Sodium-Potassium Ratio<br>(Na/K) | 0.41 | 0.34 – 0.52 | $\geq \mathbf{0.60}$<br>85<sup>th</sup> percentile<br>$N = 25$ |

### 3.2 Associations of MEP with Mother-Infant Characteristics

Demographic and selected health characteristics of the 162 participants are presented in **Table 3**. There was a weak negative association between maternal age and Na, *r*(160) = −.19, *p <* .05, and the Na/K ratio, *r*(160) = −.18, *p <* .05. Human milk Na levels also declined with older infant age, *r*(160) = −0.51, *p* < .001, but this association was not observed for the Na/K ratio.

**Table 3.** Means (SDs) or *N* (%) for maternal-infant characteristics for the entire sample (*N* = 162).

|  | Whole Sample |  |
| --- | --- | --- |
|  | N | Mean (SD)<br>Or<br>N (%) |
| Maternal Age | 162 | 31.2 (5.4) |
| Parity (Multipara) | 162 | 88 (54%) |
| Race (Black) | 162 | 98 (61%) |
| BMI (kg/m<sup>2</sup>) | 162 | 27.1 (6.5) |
| Menses Since Birth (yes) | 159 | 61 (38%) |
| Hours Since Pumping | 123 | 3.1 (2.5) |
| Infant Age (weeks) | 162 | 27.3 (19.0) |

Maternal and infant characteristics were evaluated in multivariable logistic regression models predicting elevated MEP. Infant age and maternal race were significant predictors in the model predicting elevated MEP as indicated by Na ≥ 10 mmol/L (**Table 4**). For every additional week of infant age, the odds of elevated MEP as defined by Na declined by 4%. Black women had a higher likelihood of experiencing elevated MEP by 3.3 times. BMI over 30 was not associated with elevated MEP as indicated by Na ≥ 10 mmol/L. No maternal or infant characteristics, including race or BMI, were significant in the model predicting elevated MEP as defined by the Na/K ratio ≥ 0.6.

**Table 4.** Multivariable logistic regression models and 95% confidence intervals predicting elevated mammary epithelium permeability (MEP) as indicated by Na ≥ 10 mmol/L.

|  | OR (95% CI) |
| --- | --- |
| **Elevated MEP** |  |
| **Na ≥ 10 mmol/L (*n* = 162)** |  |
| Infant Age (weeks) | **0.96 (0.93 – 0.99)** |
| Race (Black)<sup>a</sup> | **3.35 (1.03 – 10.82)** |

Model 1: <sup>a</sup>Reference: White Note: Bolded values indicate a significant finding.

### 3.3 Human Milk Cytokines and Associations with MEP

Descriptive statistics for the human milk cytokines are presented in **Table 5**. Both Na and the Na/K ratio had several significant positive associations with the selected cytokines. Nine analytes each were positively correlated with Na or the Na/K ratio, although the specific analytes varied by MEP indicator (see **Table 5**). Only leptin was not associated with either Na or the Na/K ratio. The overall pattern suggested that Na was more strongly associated with growth factors, while the Na/K ratio was more strongly associated with pro-inflammatory cytokines.

**Table 5.** Descriptive statistics for pro-inflammatory cytokines, growth factors and adipokines and Spearman rank correlations with Na and the Na/K ratio.

|  | N | Median<br>(IQR) | Na | Na/K Ratio |
| --- | --- | --- | --- | --- |
| **Pro-inflammatory cytokines** |  |  |  |  |
| IL-6 | 162 | 1.53<br>(0.45 - 3.74) | 0.35*** | 0.36*** |
| IL-8 | 157 | 249.93<br>(132.46 – 629.40) | 0.13 | 0.31*** |
| TNFα | 162 | .95<br>(0.46 – 2.51) | 0.33*** | 0.28*** |
| IL-1β | 162 | .70<br>(0.27 – 1.81) | 0.15 | 0.28*** |
| **Growth Factors** |  |  |  |  |
| FASL | 162 | 43.23<br>(34.17 – 57.15) | 0.28*** | 0.18* |
| VEGFD | 161 | 275.92<br>(209.91 – 409.83) | 0.25** | 0.11 |
| FLT1 | 161 | 1671.69<br>(1363.21 – 2423.49) | 0.24** | 0.12 |
| PLGF | 161 | 72.30<br>(41.77 – 127.57) | 0.24** | 0.26*** |
| bFGF | 162 | .98<br>(0.57 – 1.76) | 0.41*** | 0.33*** |
| EGF | 162 | 4294.31<br>(3010.36 – 6052.75) | 0.20* | 0.22** |
| **Adipokines** |  |  |  |  |
| Leptin | 162 | 1056.22<br>(490.75 – 2190.75) | 0.11 | 0.07 |
| Adiponectin | 162 | 20.80<br>(15.70 – 28.46) | 0.28*** | 0.17* |

\* $p < 0.05$ , \*\* $p < 0.01$ , $p < 0.001$

Principal components analysis, performed to consolidate the analytes, identified three unique factors (see **Table 6**). IL-6, IL-8, TNF-α, IL-1β, and EGF loaded onto a factor representing the pro-inflammatory signature. The second factor was composed primarily of cytokines involved in tissue growth and remodeling and included FASL, VEGFD, FLT1, PLGF, bFGF, and adiponectin. A single analyte, leptin, loaded onto the third factor. Pro-inflammatory and tissue remodeling factor scores were calculated based on this analysis.

**Table 6.**
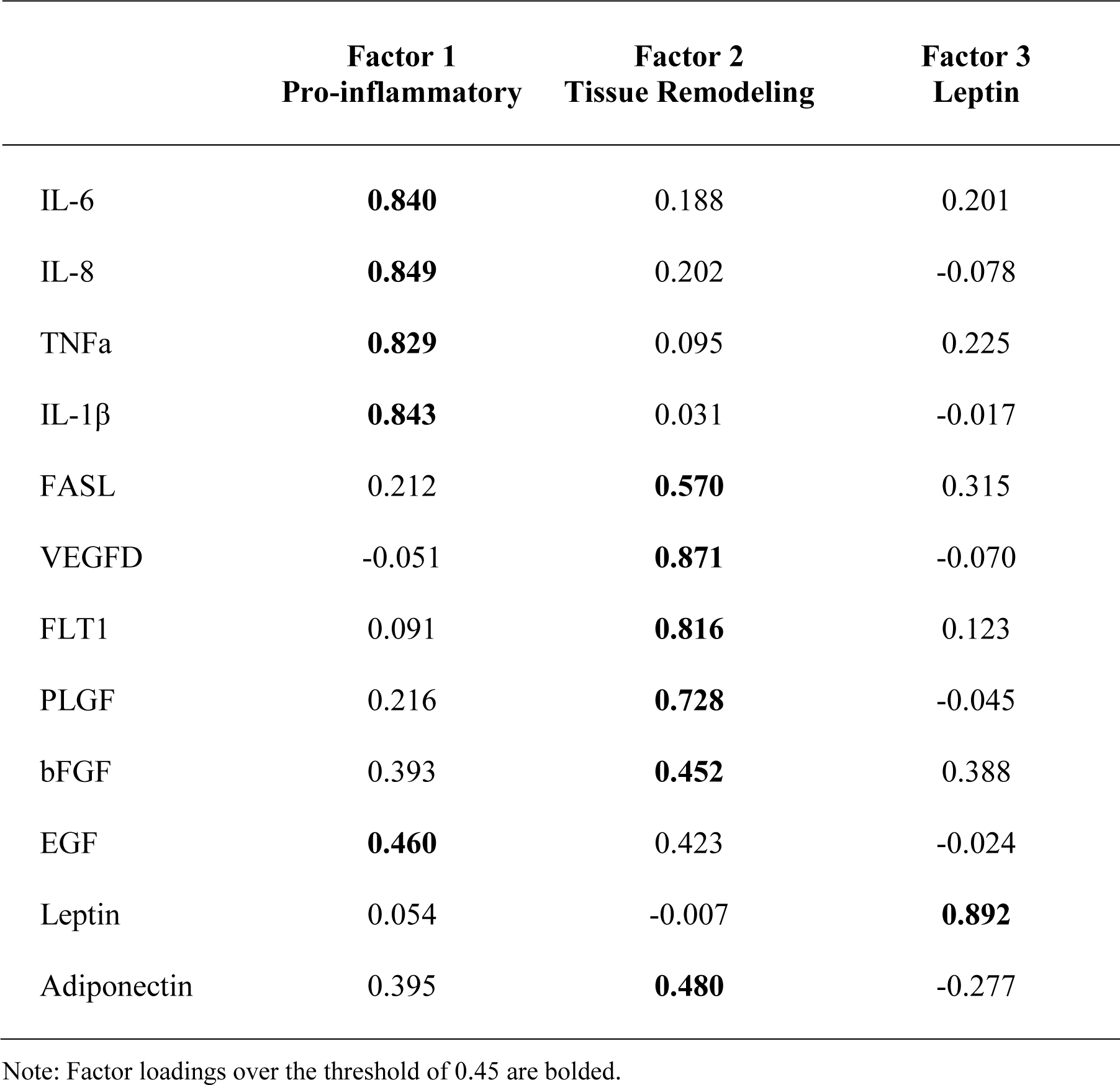
Analyte factor loadings for principal component analysis with varimax rotation.

Pro-inflammatory factor scores were higher in primipara as compared to multipara, Mann-Whitney *U =* 2391, *p* < 0.05. The pro-inflammatory factor was more strongly correlated with Na/K, *r*(154) = 0.31, *p* < 0.001, than with Na alone, *r*(154) = 0.22, *p* < 0.01. In contrast, the tissue remodeling factor was more strongly correlated with Na, *r*(154) = 0.32, *p* < 0.001, than with the Na/K ratio, *r*(154) = 0.19, *p* < 0.05.

Multivariable logistic regression models were examined to evaluate pro-inflammatory and tissue remodeling factor scores as predictors of elevated MEP (**Table 7**). Controlling for infant age and maternal race, higher pro-inflammatory and tissue remodeling factor scores each uniquely raised the odds of elevated MEP as indicated by Na ≥ 10 mmol/L. A similar pattern was observed for the Na/K ratio where pro-inflammatory and tissue remodeling factors were uniquely associated with elevated MEP during established lactation.

**Table 7.** Multivariable logistic regression models and 95% confidence intervals predicting increased mammary epithelium permeability (MEP) from pro-inflammatory and tissue remodeling factor scores.

|  | OR (95% CI) |
| --- | --- |
| **Elevated MEP** |  |
| **Na ≥ 10 mmol/L ($n = 156$)** |  |
| Infant Age (weeks) | 0.97 (0.93 – 1.00) |
| Race (Black)<sup>a</sup> | 3.90 (0.98 – 15.54) |
| Pro-inflammatory Factor | **2.40 (1.39 – 4.17)** |
| Tissue Remodeling Factor | **2.55 (1.42 – 4.57)** |
| **Elevated MEP** |  |
| **Na/K ≥ 0.6 ($n = 156$)** |  |
| Pro-inflammatory Factor | **2.45 (1.51 – 4.00)** |
| Tissue Remodeling Factor | **2.02 (1.25 – 3.26)** |

Model 1: <sup>a</sup>Reference: White
Notes: Bolded values indicate significant associations.

## 4 Discussion

To our knowledge, this is the first study to examine associations between MEP and cytokine profiles in human milk during *established lactation* among women living in the US. Of note, the percentage of participants with elevated MEP was higher in this cohort than was observed in a European cohort during established lactation using the same criteria (Na/K > 0.6) (27). Elevated MEP during established lactation occurred in 5% or less of women in a European cohort as compared to the observed 15% in the current study.

Historically, MEP has been studied primarily in relation to secretory activation, subclinical mastitis (SCM), mastitis, and involution. Secretory activation occurs in the first days postpartum and is associated with a precipitous decline in MEP (17,20), while SCM and mastitis may occur at any time during lactation and are associated with significant increases in MEP (2,27–30). The milk samples assessed in the present study were all from participants nursing infants between 1 and 18 months of age, and to our knowledge, no participant was experiencing symptoms of mastitis at the time of milk collection. While we did not specifically inquire about plans for weaning, it is possible that some participants may have been in the midst of this process. MEP, as indicated by elevated Na or Na/K, is known to increase during involution (31). Of note, the odds of elevated MEP (defined as Na > 10 mmol/L) declined with advancing infant age overall, possibly representing normal changes in mammary function over the course of the lactation cycle.

As predicted, Black participants in our cohort were more likely to experience elevated MEP (defined as Na > 10 mmol/L). Since this was the first study to assess MEP in a sizable (*n* = 98) sample of Black women in the US, it is unknown whether the 3.3 times greater odds of elevated MEP among Black women was due to socioeconomic conditions impacting the frequency of breastfeeding or pumping, represents normal variation in healthy breast tissue, or was a sign of inflammation in the mammary gland. Elevated MEP among Black women is consistent with our previous report demonstrating higher levels of some pro-inflammatory cytokines in the milk of Black women in this cohort (24).

Levels of all cytokines examined were positively associated with either Na or the Na/K ratio, with the exception of leptin. A body of research has previously identified links between cytokines and permeability. Many of these studies have suggested that inflammation may drive increased permeability (2,4,29,30). Indeed, during mastitis open paracellular pathways may be adaptive, allowing cytokine-producing leukocytes access to the alveolar lumen (32). However, permeability may also increase in response to the rising alveolar pressure associated with milk accumulation. Eventually, this can lead to tissue remodeling as seen during involution. Both pro-inflammatory cytokines and tissue growth factors play a role in this process (14,18).

In the current study, pro-inflammatory and tissue remodeling factors each uniquely raised the odds of experiencing elevated MEP as indicated by either Na ≥ 10 mmol/L or the Na/K ratio ≥ 0.6. However, when considering continuous measures of Na or the Na/K ratio, a pattern emerged where the pro-inflammatory factor was more strongly associated with the Na/K ratio, while the tissue remodeling factor was more tightly linked with Na. To our knowledge, this is the first study to examine growth factors in relation to MEP. While additional research is needed to confirm this finding, unique patterns of Na and Na/K with specific cytokines could be used to identify the physiologic processes underlying elevated MEP.

Identifying the etiology underlying MEP has important implications for both parent and infant. Elevated MEP has been linked to delayed onset of lactation (17), low milk supply (33), reduced milk nutrient content (27), and inadequate infant growth (4). Infant growth faltering has also been identified in relation to elevated cytokines in human milk (3). Growth factors in human milk may also affect the development of the infant intestinal barrier (11). Identifying persistently increased permeability could also have important implications for identifying breast cancer risk, given the established role of cytokines in tumorigenesis (9,24,34). Taken together, both MEP and cytokines could have important implications for both parent and infant health.

An important direction for future research is to determine how to most appropriately measure MEP. In the present study, analyses based on Na levels and Na/K ratios provide slightly different results. Using cut-off values from the literature (17), Na was the more conservative indicator of elevated MEP in this study, identifying 19 cases as compared to the 25 cases identified using the Na/K ratio.

Of note, a subset of 17 cases was identified by both indicators, while 2 cases were identified by Na alone, and 8 cases by the Na/K ratio alone. As noted above, continuous measures of Na were also more closely associated with tissue growth and remodeling cytokines, while the Na/K ratio seemed to be more tightly linked with pro-inflammatory cytokines. Additional research is needed to determine how best to interpret human milk Na levels and Na/K ratios during established lactation.

There were several strengths to the current study including a racially diverse cohort, representation of women up to 18 months of established lactation, and a panel of 12 cytokines. However, there is also an important limitation. This secondary analysis used milk and questionnaire data from a study aimed at understanding factors associated with breast cancer risk. Therefore, the study design and questionnaires were not optimally structured to assess factors known to be associated with MEP.

## 5 Conclusions

Results presented here highlight the importance of measuring mammary epithelium permeability in studies of normal developmental and inflammatory processes in the human mammary gland, as well as in studies of the effects of human milk on infant health. Surprisingly, the human mammary gland is the only organ for which we lack routine clinical tests for normal function (35,36). Rich information may be obtained through the measurement of MEP indicators during established lactation. Research aimed at understanding the importance of MEP for mammary gland health or infant growth and development should be prioritized.

## Data Availability

All data produced in the present study are available upon reasonable request to the authors.

## Author Contributions

Katie Kivlighan: conceptualization, formal analysis, writing – original draft

Sallie Schneider: conceptualization, methodology, writing – review & editing

Eva Browne: methodology, validation, investigation, data curation, review

Brian Pentecost: writing – review & editing

Douglas Anderton: writing – review & editing

Kathleen Arcaro: conceptualization, methodology, investigation, resources, supervision, visualization, writing – review & editing

## Funding

Funding was provided by the College of Natural Sciences, University of Massachusetts-Amherst.

### Acknowledgments

We wish to thank the participants who donated milk for this research.

